# Magnitude, pattern and correlates of multimorbidity among patients attending chronic outpatient medical care in Bahir Dar, northwest Ethiopia: the application of latent class analysis model

**DOI:** 10.1101/2021.10.08.21264745

**Authors:** Fantu Abebe Eyowas, Marguerite Schneider, Shitaye Alemu, Sanghamitra Pati, Fentie Ambaw Getahun

## Abstract

**Objective:** This study aimed to investigate the magnitude, pattern and associated factors of multimorbidity in Bahir Dar, Ethiopia.

**Methods:** A multi-centered facility based study was conducted among 1440 participants aged 40+ years attending chronic outpatient medical care. Two complimentary methods (interview and review of medical records) were employed to collect the data on socio-demographic, behavioral and disease related characteristics. The data were analyzed by STATA V.16 and R Software V.4.1.0. We run descriptive statistics and fitted logistic regression and latent class analyses (LCA) models to determine associated factors and patterns of multimorbidity. Statistical significance was considered at p-value ≤0.05.

**Results:** The magnitude of individual chronic conditions ranged from 1.4% to 37.9%, and multimorbidity was identified in 54.8% (95% CI=52.2%-57.4%) of the sample. The likelihood of developing multimorbidity was higher among participants aged from 45-54 years (AOR: 1.5, 95%CI= 1.1, 2.1), 55-64 years (AOR: 2.5, 95%CI=1.7, 3.5) and 65 years or more (AOR: 2.4, 95%CI=1.7, 3.5), among individuals classified as overweight (AOR: 1.6, 95%CI=1.2, 2.1) or obese (AOR: 1.9, 95%CI=1.3, 3.0) and among those individuals who believe in external locus of control (AOR: 1.8, 95%CI=1.3, 2.5). Four patterns of multimorbidity were identified, the cardiovascular category being the largest class (50.2%), followed by the metabolic group (32.6%). Advanced age, overweight and obesity predicted latent class membership, adjusting for relevant confounding factors.

**Conclusion:** The magnitude of multimorbidity in this study was high. The most frequently diagnosed chronic conditions shaped the patterns of multimorbidity. Advanced age, overweight and obesity were the factors profoundly associated with multimorbidity. Health service organization and provision in the study area need to be oriented by the realities in disease burden and pattern of multimorbidity. Further research is required to better understand the impact of multimorbidity on individuals wellbeing, survival and health service delivery.

## Background

Globally, one in four adults are living with at least one chronic conditions, of which 80% of them have two or more long lasting and incurable non-communicable diseases (NCDs), commonly knowns as multimorbidity (1). Multimorbidity is a relatively new concept in research globally (2) and there remains a huge interest in answering what is philosophically adequate to define the concept (3) and measure its epidemiology (4, 5). Further, there is no universally accepted standard for what constitute chronic conditions and how many chronic conditions to consider towards the measurement of multimorbidity (6, 7).

This challenge led researchers to employ quite diverse methodologies to study the epidemiology of multimorbidity in their context (7, 8). Inconsistencies were mainly observed in terms of the age group of the population studied, the types and number of chronic NCDs considered, study setting, method of data collection and sample size, among others (9, 10). The use of different methodologies resulted in a huge variation in the prevalence estimates and difficulty in comparing and pooling results of multimorbidity studies globally (6, 11).

The need to understand the patterns of disease combinations/clusters and associated complexity and care is also well recognized (12). Certain conditions may be more likely to cluster than others (13). This may be the consequence of shared pathophysiological pathways, in which the presence of one condition increases the risk of another, or may reflect common environmental or biological risk factors (10, 14). However, the available literature is not consistent in the methodologies to determine the patterns of multimorbidity and the factors underlying the patterns of multimorbidity (15).

Another issue with the current definitions of multimorbidity is that, they take little or no account of how people themselves define their problems and too little attention is paid to what matters most to individuals living with multiple health problems (16).

Despite the inconsistency in the methodologies employed to define and measure multimorbidity, however, evidence has shown that the burden of multimorbidity is growing globally(2, 17). Highest prevalence rates were reported from studies in high income countries, where about one in four adults experience multimorbidity (16, 18). A few studies in low-and middle-income countries (LMICs) have also reported an increasing trend in the burden of NCDs multimorbidity (2, 17, 19).

The prevalence of multimorbidity increases substantially with age (6). However, the absolute number of people with multimorbidity has also been found to be higher in those younger than 65 years (1, 10). Multimorbidity is also associated with socioeconomic deprivation and evidence has shown that those living in the most deprived areas develop multimorbidity 10 to 15 years earlier than in the least deprived decile of the population and tend to have multimorbidity accompanying mental health problems (1). Female gender and obesity are also common risk factors for the occurrence of multimorbidity globally (9). It is argued that patients with multimorbidity are more than the sum of individual conditions (20).

Compared to patients without multimorbidity, those living with multimorbidity have a higher mortality rate, worse health related quality of life (HRQoL), impaired functionality, higher rate of hospital admission and related health and social care costs (16, 18).

People living with multimorbidity need more holistic, generalist long-term care and support than patients having a single NCD (21, 22). However, most patients with multiple chronic conditions may have more than one physician, such as one from each relevant specialty often working in silos and are prescribed with more drugs (polypharmacy) for long periods of time often leading to dangerous drug to drug or drug to diseases interactions and other life threatening complications (23). They also face challenges in navigating the health care system and managing their health, and are generally less satisfied with the care they receive (16). Further, the rapid emergence of infections such as COVID-19 are fueling the complexity and posing a huge burden to the health systems and worsening outcomes of patients with preexisting chronic diseases and multimorbidity (24, 25).

The impact of multimorbidity is likely to be significant in LMICs, including Ethiopia where health systems are overwhelmed by the high speed of NCD growth and high burden of communicable diseases such as HIV and Tuberculosis (26). On the other hand, health systems in LIMCs are largely configured with conventional one-size-fits-all chronic care model (18), and access to NCDs care is inadequate to the poor, furthering disease accumulation and long-term financial burden (16). Therefore, it is likely that patients with multimorbidity face accumulating and overwhelming complexity resulting from the sum of uncoordinated responses to each of their health problems (27-29).

Despite the huge challenge multimorbidity might pose, there is a significant knowledge gap in terms of the burden, associated risk factors, and the pattern and distribution of multimorbidity in Ethiopia. If health systems are to meet the needs and priorities of individuals living with multimorbidity in Ethiopia, we must first accurately measure the burden and distribution of NCDs multimorbidity in the context.

This study aimed to employ a blend of methods to determine the magnitude and pattern of multimorbidity and associated factors among individuals attending chronic outpatient medical care in public and private health facilities in Bahir Dar city, Ethiopia. We conducted the study as planned and revealed valuable findings.

The findings of this study will contribute to the sparse evidence on the burden and pattern of multimorbidity and risk factors associated in LMICs. The study will also offer valuable insight to researchers, policy makers and practitioners for shaping future research endeavors and devise preventive and management strategies vis-a-vis multimorbidity of NCDs in the country and beyond.

## Methods

This is a multi-center facility based study conducted in public and private health facilities rendering health services in Bahir Dar City, Ethiopia. The city is the capital of Amhara region-the second populous region in the country where about 30 million people are living. The detail of the methods employed in this study has been published elsewhere (30) and it is summarized as follows.

### Study setting and population

This study was conducted in five hospitals (three public and two private) and three private higher/specialty clinics in Bahir Dar city, north-west Ethiopia. These facilities provide the bulk (∼80%) of chronic NCDs care for the people living in the city and surrounding residences. Chronic NCDs care and management is presumed to be provided in a relatively uniform fashion using the national NCDs treatment guideline (31). However, the nature of patients vising these facilities may vary and there remains a concern on quality and affordability NCDs care in public hospitals and private health facilities, respectively.

A two-stage clustered stratified random sampling method was adopted for recruiting facilities and participants. The sample size from each facility has been determined based on the notion of probability proportional to size (PPS) using the pool of chronic NCD patients (≥ 40yrs) registered for follow-up over the year preceding our assessment (January -December 2020) in each participating facility.

Only facilities who were providing chronic NCDs care by general practitioners or specialist physicians for at least a duration of one year prior to the data collection period were considered. Older adults (40 years or more) diagnosed with at least one NCD and are on chronic diseases follow up care for at least six months prior to the study period were recruited for the study. Pregnant women and individuals who are too ill to be interviewed and admitted patients were excluded.

A sample of 1440 adult patients (≥40yrs) attending chronic outpatient medical care in the selected health facilities were randomly selected and enrolled from March 15 to April 30, 2021 for this baseline study.

### Definition and measurement of Multimorbidity

Multimorbidity was operationalized as the co-occurrence of two or more of the chronic NCDs specified below. List of NCDs considered in this study were determined based on a review study (30), and includes hypertension, diabetes, heart diseases (heart failure, angina and heart attack), stroke, bronchial asthma, chronic obstructive pulmonary diseases (COPD), depression and cancer.

Moreover, based on a pilot study we conducted, other prevalent (accounting 2-5%) chronic diseases, including musculoskeletal disorders (arthritis, chronic back pain and osteoporosis), thyroid disorders (hyperthyroidism and multinodular goiter), chronic kidney disease, gastrointestinal disorders (chronic liver, gall bladder and gastric diseases) and Parkinson’s disease (PD) were identified and included in our assessment. Information on these chronic conditions was assessed through a question about ever being diagnosed with the disease by a health professional. The specific question was, “have you ever been told by a health professional/doctor that you have (disease name)?” responses were either yes (scored as “1”) or no (score as “0”). Participants were also prompted to report up to three additional chronic conditions they are living with if any.

To improve the quality of data obtained from interviews (32, 33), we reviewed medical records of all the study participants. Direct assessment of chronic diseases was not, however, possible due to resource constraint and methodological challenges.

### Measurement of Independent variables

Independent variables including socio-demographic characteristics [age, gender, education, wealth index, marital status, family size, residence and occupation], dietary habits [amount and frequency of fruit and vegetables consumption and amount of daily salt consumption], behavioral and lifestyle patterns [alcohol consumption, smoking, Khat consumption, physical exercise], patient activation (PA) status, social support system and locus of control (LoC) were measured by interview using validated tools. Whereas, data to calculate body mass index (BMI) and waist and hip circumference were directly measured from patients according to the approaches described below.

Height and weight was measured using standardized techniques with participants barefoot and wearing light clothing. Participants height was measured to the nearest 0.1 cm using a portable Seca 213 Stadiometer and weight was recorded to the nearest 0.1 kg using a weighing scale. These data were used to calculate individual body mass index (BMI; kg/m^2^). BMI values are classified into categories for each individual based on established WHO cut-offs for BMI, which included four categories: underweight (<18.5 kg/m^2^), normal (18.5–24.9 kg/m^2^), overweight (25.0–29.9 kg/m^2^), and obese (30 kg/m^2^)(34).

A flexible, stretch-resistant tape was used to measure waist and hip circumference to the nearest 0.1 cm midway between the 12^th^ rib and the iliac crest and around the widest portion of the hips, respectively. Participants waist-to-hip ratio (WHR) was estimated and interpreted according to the WHO’s protocol (35).

Patient activation (PA) was assessed using validated tools (36, 37). The tool contains 13 statements answered on a 4-point Likert-type scale about managing one’s health and summed to a 100-point scale, with higher scores reflecting higher levels of activation (38). The score was classified into four stages, the lowest category being poor activation (≤47.0 = stage 1, 47.1-55.1=stage 2, 55.2-67.0=stage 3 and ≥67.1=stage 4).

Social networking and support system was assessed through face-to-face interview using pre-tested and standardized tools (Oslo Scale) (39). A scale ranging from 3–8 was interpreted as poor social support, 9– 11 moderate social support and 12–14 strong social support. Multidimensional health locus of control scale (form C) was used to assess health-related control beliefs (locus of control) of the people living with chronic NCDs (40). The 18-item scale was scored using Likert scale as strongly agree (6 points) to strongly disagree (1point).

Wealth Index (a latent construct) at household level was generated from a combination of material assets and housing characteristics (41). The Wealth index was scored using principal component analysis (PCA) technique. The score was classified into quintiles, differently for urban and rural residents separately, while quintile 1 represents the poorest and quintile 5 the wealthiest (42). It was collapsed into three classes as low, middle and high income for simplifying its interpretability.

### Data Collection Tools and Procedures

A range of standardized tools, including the WHO’s STEP survey questionnaire were translated and pilot tested and used for the data collection. For the sake of a more efficient and accurate data collection, aggregation and statistical analysis, the data were collected by the Kobo Toolbox software(43). Patients were interviewed and assessed following consultation periods. Physicians and nurses working in the chronic care unit were involved in the data collection process. However, data were primarily collected by ten graduate nurses recruited from institutions outside the study facilities.

After obtaining consent from the participants, information on socio-demographic characteristics, dietary practices, lifestyle habits, doctor diagnosed medical condition/s (specified above), activation status (PA), psychosocial support, locus of control and depression level was collected by face-to-face interview. Then, measurement of weight, height and waist circumference was made. Finally, patient charts (medical records) were reviewed to capture recorded medical diagnoses, medications prescribed and fasting blood glucose (FBG) and glycated hemoglobin (HbA1c) values and HIV status if available. In addition, data on COVID-19 infection was gathered at different point in time through patient interview and review of medical records.

### Data Quality Assurance

Data were collected from multiple sources using pilot tested and standardized instruments. That helped us to validate and rectify congruence between the data captured from different sources (interview and review of medical records). The use of Kobo toolbox software helped to collect real time data and monitor the validity of the information entered daily from all the data collector (43). The questionnaires to collect the data were translated to Amharic (local language) and pilot tested for cross-cultural adaptability based on standard protocols (44, 45). Data collectors and supervisors received a two-days high level of training detailing the study, including obtaining written consent, conducting face-to-face interview, performing physical measurement, medical record review and navigating through the questionnaires in the Kobo toolbox platform preloaded into their smart phones. The data collection process was monitored by trained supervisors and the principal investigator and the data sent every day to the Kobo toolbox server were checked for completeness, accuracy and clarity.

### Data Analysis

The data from Kobo toolbox was downloaded into excel spreadsheet and exported to SPSS V. 21 for cleaning and finally analyzed by STATA V. 16 and R V.4.1.0. Descriptive statistics were computed to describe the sociodemographic, lifestyle and other characteristics of participants.

The magnitude of individual chronic conditions was determined and Chi-squared test was run to determine the difference in distribution of individual NCDs between men and women.

The prevalence of multimorbidity was determined by combining evidences mainly from two sources, patient interview and medical record review. The dependent variable (multimorbidity) was coded as yes and no.

Association between the dependent variable (multimorbidity) and independent variables was assessed by fitting bivariate logistic regression model and the unadjusted odds ratio (OR) with 95% confidence intervals and p-values are reported for each of the independent variable analyzed. Variable having a p-vale <0.2 were fitted into the multivariable logistic regression model to predict the adjusted effect of the independent variables on multimorbidity. Before running the multivariable analysis multi-collinearity between independent variables was checked using the Variance Inflation Factor (VIF), and variables were not strongly correlated (the highest value was 2.11). A p – value ≤ 0.05 was taken as statistically significant.

Latent class analysis (LCA)(46) was fitted into R 4.1.0 software (poLCA package) to identify the subgroups of patients sharing class membership. LCA is a powerful method for identifying unobserved groups with categorical outcome variables such as patterns of multimorbidity in a given population(47). Identifying subgroups of individuals could help designing appropriate interventions in the pursuit of multimorbidity (48). Given that there is no single indicator reflecting an optimal model fit, model selection was based on a balance of parsimony, substantive consideration, and several fit indices(49).

We used the Akaike Information Criterion (AIC) and the Bayesian Information Criterion (BIC) to evaluate the relative fit and adequacy of the models. Lower values on the AIC and the BIC indicate a better-fitting model(49). The BIC tends to select simpler models than the AIC, and in a Monte Carlo simulation it has been shown to be the most reliable criteria when deciding on the optimal latent class model [39].

Predictors of class membership were identified by running multinomial logistic regression analysis. All the assumptions for multivariable and multinomial logistic regression analyses were checked and the fit statistics reported accordingly. Key parameter estimates for fitting an adequate LCA model were also assessed and reported accordingly.

## Results

### Characteristics of the Study Participants

Of the total 1440 study participants enrolled, complete data were obtained from 1432 individuals giving rise to a response rate of 99.4%. The majority (n=1007, 70.3%) of study participants were urban residents and females constitute a slightly higher (51%) percentage in terms of sex distribution. The mean age of the participants was 56.4 (ranged from 40-93 years) with a standard deviation (SD) of 11.8 years. Individuals aged 45-54 years and 55-64 years accounted equally (27.9) for the age distribution and those aged 65+has 26.9% share from the total sample (Table 1).

**Table 1:** Socio-demographic characteristics of study participants attending chronic outpatient NCDs care in Bahir Dar, Ethiopia (N=1432)

| Variables | Frequency | Percentage |
| --- | --- | --- |
| Age |  |  |
| ≤44Yrs | 247 | 17.3 |
| 45-54Yrs | 399 | 27.9 |
| 55-64Yrs | 400 | 27.9 |
| 65+Yrs | 386 | 26.9 |
| Sex |  |  |
| Male | 702 | 49.0 |
| Female | 730 | 51.0 |
| Marital status |  |  |
| Currently married | 1081 | 75.5 |
| Single* | 351 | 24.5 |
| Education |  |  |
| No formal education | 780 | 54.5 |
| Primary | 166 | 11.6 |
| Secondary | 171 | 11.9 |
| College level and above | 315 | 22.0 |
| Occupation |  |  |
| Housewife | 329 | 23.0 |
| Employed | 328 | 22.9 |
| Farmer | 288 | 20.1 |
| Trader | 207 | 14.5 |
| Retired | 141 | 9.8 |
| Unemployed | 139 | 9.7 |
| Wealth Index (SES) |  |  |
| Poorest | 269 | 18.8 |
| Poorer | 334 | 23.3 |
| Middle | 267 | 18.6 |
| Rich | 252 | 17.6 |
| Richest | 310 | 21.6 |

\*Includes never married, divorced, widowed and separated

The majority of them (n=1081, 75.5%) were married at the time of data collection. Looking into education level of the respondents, the majority (n=780, 54.5%) did not attend any formal education. While 11.6%, 11.9% and 22% of the participants attended primary, secondary and college level or above education programs, respectively. Housewives (23%) and employed individuals (22.9%) represent the largest share in the occupation category. The highest percentage (n=536, 37.4%) of the participants had low SES, and 28.3% (n=405) and 34.3% (n=491) of them fell on the middle and highest SES category, respectively (Table 1).

### Lifestyle, Behavioral and Psychosocial Characteristics

Thirty-seven percent (n=530) of the participants reported that they were doing moderate/high intensity exercise at least three times per week. With regard to dietary habits, 563 (39.3%) and 737 (51.5%) reported to have consumed fruit and vegetables three times or more per week, respectively. About one sixth (n=220, 15.4%) of them reported to have taken alcohol of any kind in the past one week prior to the interview and 89 (6.2%) of the participants have had past history of smoking.

The highest percentage (n=763, 53.3%) of participants had normal body mass index (BMI), whereas overweight and obesity together accounted 32%. However, based on the waist-to-hip ratio health risk classification, the highest proportion (n=810, 56.6%) were on high health risk group.

With regard to social support and networking with people, the majority (n=726, 50.7%) reported that they have moderate social support, and about one third (28%) reported strong social support and the remaining 21% reported that they have a poor social support. The highest percentage of the participants believe that life events including illnesses are due to chance (n=1245, 86.9%) (Table 2).

**Table 2:** Lifestyle, behavioral and psychosocial characteristics, of participants attending chronic outpatient NCDs care in Bahir Dar, Ethiopia (N=1432)

| Variables | Frequency | Percentage |
| --- | --- | --- |
| Moderate/High intensity physical exercise |  |  |
| Yes | 530 | 37.0 |
| No | 902 | 63.0 |
| Fruit consumption |  |  |
| Yes 3 or more times/week | 306 | 21.4 |
| Yes 1-2 times/week | 256 | 17.9 |
| No/<1 consumption/week | 870 | 60.7 |
| Vegetables consumption |  |  |
| Yes 3 or more times/week | 698 | 48.7 |
| Yes 1-2 times/week | 262 | 18.3 |
| No/<1 consumption/week | 472 | 33.0 |
| Smoking (Past history) |  |  |
| Yes | 89 | 6.2 |
| No | 1343 | 93.8 |
| Alcohol consumption |  |  |
| Yes | 220 | 15.4 |
| No | 1212 | 84.6 |
| Khat Consumption |  |  |
| Yes | 40 | 2.8 |
| No | 1392 | 97.2 |
| BMI |  |  |
| <18.5 kgm2 | 211 | 14.7 |
| 18.5-24.9 kgm2 | 763 | 53.3 |
| 25.0-29.9 kgm2 | 326 | 22.8 |
| ≥30 kgm2 | 132 | 9.2 |
| Waist-to-hip ratio |  |  |
| Low health risk | 399 | 27.9 |
| Moderate health risk | 223 | 15.6 |
| High health risk | 810 | 56.6 |
| Dyslipidemia |  |  |
| Yes | 54 | 0 |
| No | 731 | 647 |
| Internal Locus of control (mean=5.2) |  |  |
| Low | 846 | 59.1 |
| High | 586 | 40.9 |
| Locus of control (chance) (mean= 3.8) |  |  |
| Low | 187 | 13.1 |
| High | 1245 | 86.9 |
| Locus of control (powerful others) (mean=4.2) |  |  |
| Low | 870 | 60.8 |
| High | 563 | 39.2 |
| HIV status |  |  |
| HIV Positive | 53 | 3.7 |
| HIV negative | 183 | 12.8 |
| Status unknown | 1196 | 83.5 |

**Table 3:** Sex Distribution of NCDs among people attending chronic outpatient NCD care in Bahir Dar, Ethiopia (N=1432)

| Type of NCD | Males | Females | Total | P. Value |
| --- | --- | --- | --- | --- |
|  | Frequency (%) | Frequency (%) | Frequency (%) |  |
| Hypertension | 453 (49.8) | 457 (50.2) | 910 (63.5) | 0.475 |
| Diabetes | 323 (53.0) | 286 (47.0) | 609 (42.5) | **0.010** |
| Heart diseases♣ | 173 (47.3) | 193 (52.7) | 366 (25.6) | 0.467 |
| Stroke | 74 (58.7) | 52 (41.3) | 126 (8.8) | **0.025** |
| Depression | 48 (39.0) | 75 (61.0) | 123 (8.6) | **0.023** |
| Asthma | 51 (56.7) | 39 (43.3) | 90 (6.3) | 0.157 |
| Chronic renal disease | 39 (60.0) | 26 (40.0) | 65 (4.5) | 0.076 |
| Musculoskeletal diseases | 35 (62.5) | 21 (37.5) | 56 (3.9) | **0.042** |
| Gastrointestinal diseases | 22 (52.4) | 20 (47.6) | 42 (2.9) | 0.755 |
| Thyroid disorders | 2 (6.5) | 29 (93.5) | 31 (2.2) | **<0.001** |
| COPD | 10 (35.7) | 18 (64.3) | 28 (2.0) | 0.183 |
| Degenerative nerve diseases | 8 (29.6) | 19 (70.4) | 27 (1.9) | 0.051 |
| Cancer | 9 (45.0) | 11 (55.0) | 20 (1.4) | 0.823 |

♣Includes heart failure, angina, heart attack and cardiac arrhythmias

In terms of activation status (PAM), the majority (63.5%) had a highest activation status. Whereas, the score of individual classified in stage one, stage two and stage three accounted 15.2%, 12.2% and 9.0%, respectively.

### Magnitude of individual NCDs and number of chronic NCDs per person

The magnitude of each of the chronic conditions considered in this study is shown in figure 1. The number of NCDs identified per person ranged from one to four (mean=1.74, SD=0.78). Hypertension was the most frequently reported NCD (63.5%), followed by diabetes (42.5%) and heart diseases (25.6%).

**Figure 1:**
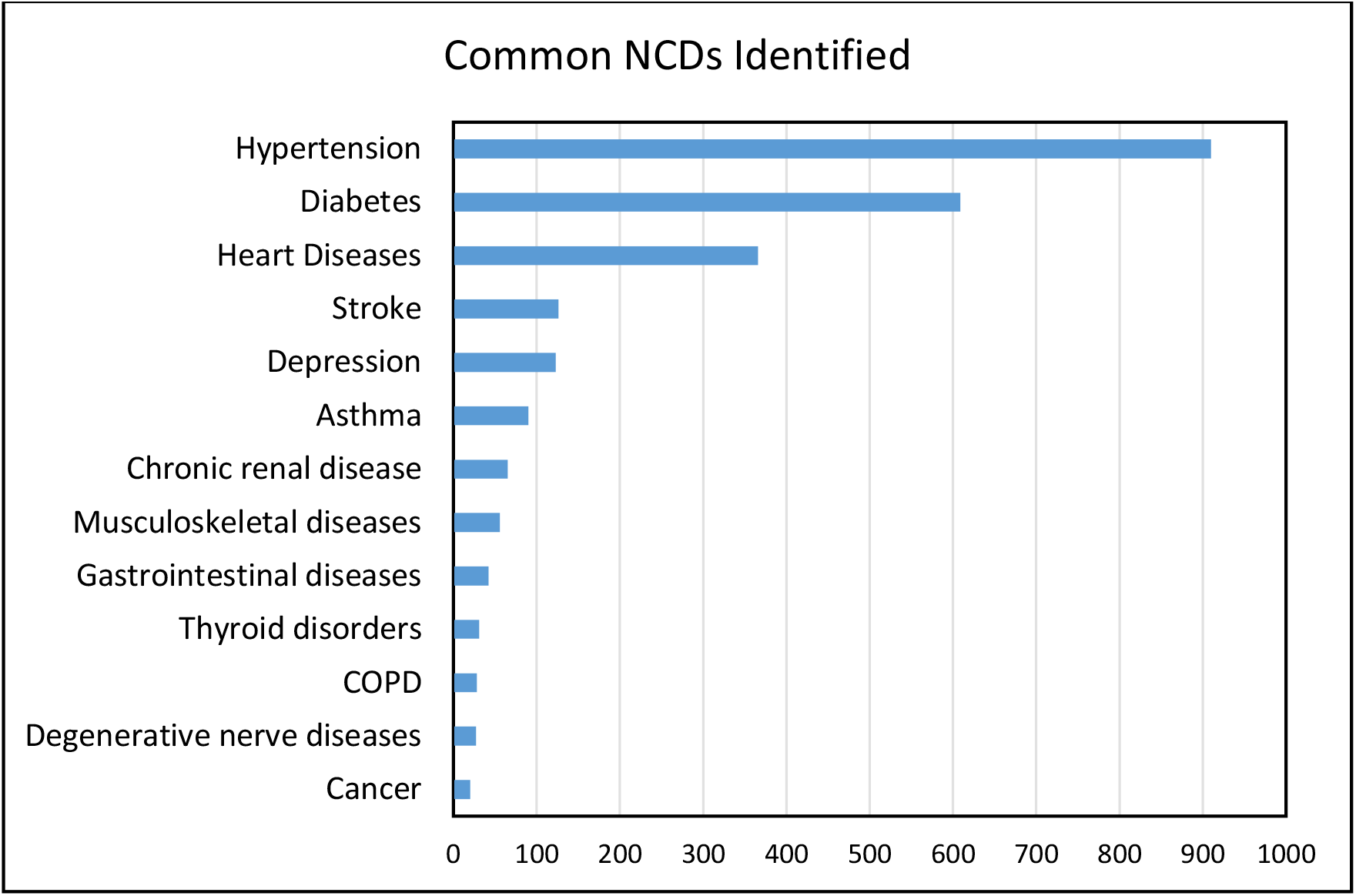
List of NCDs studied and their magnitude among participants attending chronic outpatient NCDs care, Bahir Dar, Ethiopia (N=1432)

Apparent gender differences were observed in the distribution of individual NCDs, including diabetes, stroke, depression and thyroid disorders (Table 4).

**Table 4:** Distribution of single NCDs and their pairwise and triples or quadruples combination, among people attending chronic outpatient NCD care in Bahir Dar, Ethiopia (N=1432)

| Single morbidity | Common pairs of NCDS | Common Triples of NCDs |
| --- | --- | --- |

| Chronic NCD | Frequency(%) | Combination | Frequency(%) | Combination | Frequency(%) |
| --- | --- | --- | --- | --- | --- |
| Hypertension alone | 245 (37.9) | Hypertension +Diabetes | 217 (38.2) | Hypertension +Diabetes+ heart diseases | 19 (8.7) |
| Diabetes alone | 225 (34.8) | Hypertension + Heart diseases | 108 (19.0) | Hypertension +Diabetes + depression | 18 (8.3) |
| Heart diseases alone | 120 (18.5) | Hypertension + stroke | 38 (6.7) | Hypertension +Diabetes + other NCDs | 49 (22.5) |
| All other forms of single NCDs<sup>a</sup> | 57 (8.8) | Hypertension +Musculoskeletal diseases | 23 (4.0) | Hypertension +heart diseases + other NCDs | 43 (19.7) |
|  |  | Hypertension + Asthma | 21 (3.7) | Hypertension + Diabetes + heart diseases + other NCDs | 12 (5.5) |
|  |  | Hypertension + Chronic Renal diseases | 21 (3.7) | Hypertension + Diabetes + two other NCDs | 13 (6.0) |
|  |  | Hypertension + Depression | 18 (3.2) | Hypertension + two or other NCDs | 36 (16.5) |
|  |  | Hypertension + other chronic diseases | 25 (4.4) | Diabetes + two or more other NCDs | 13 (6.0) |
|  |  | Diabetes + Depression | 8 (1.4) | Heart diseases + two or more other NCDs | 11 (5.0) |
|  |  | Diabetes + heart disease | 6 (1.0) | Triple or quadruple of all other NCDs | 4 (1.8) |
|  |  | Diabetes + other chronic NCDs | 25 (4.4) |  |  |
|  |  | Heart disease +Depression | 16 (2.8) |  |  |
|  |  | Heart diseases + other chronic diseases | 27 (4.8) |  |  |
|  |  | Comorbidity of other NCDs | 14 (2.5) |  |  |

<sup>a</sup> Includes asthma, COPD, stroke, cancer and depression

More than half 54.8% (CI=52.2%-57.4%) of the study participants had multimorbidity. Of which, 39.6% (n=567) had two NCDs and 15.2% (n=218) of them had three or more chronic NCDs. Whereas, six hundred forty-seven (45.2%) participants had single morbidity, from which hypertension (37.9%), diabetes (34.8%) and heart diseases (18.5%) were the commonest NCDs identified among individuals living with single NCD (Figure 2).

**Figure 2:**
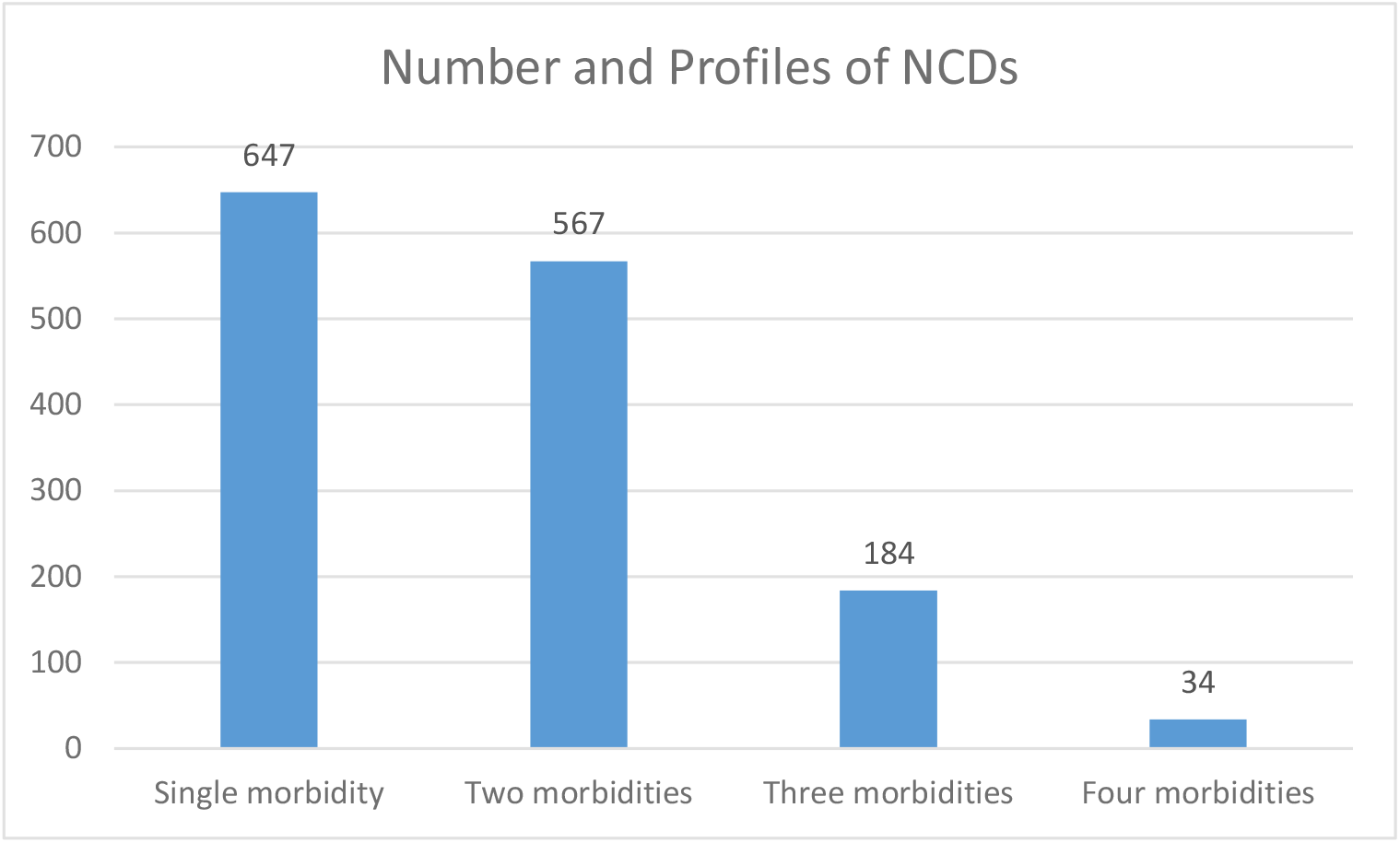
Patterns of NCDs morbidity among individuals attending chronic NCDs care in Bahir Dar, Ethiopia (N=1432)

The most prevalent NCDs have highly contributed for shaping the patterns of multimorbidity in this study: For example, hypertension was co-existed with diabetes and heart diseases in 38.2% and 19.0% of the participants, respectively. Similarly, co-occurrence of diabetes was observed among individual with heart diseases, depression and other types of reported chronic conditions. Hypertension remain the most frequently reported NCD (87.2%) among individuals living with three or more NCDs in our study. Diabetes was reported by 51% of those who had three or more chronic NCDs and heart diseases were reported by 39% of the participants from this group (Table 4).

### Factors associated with multimorbidity of chronic NCDs

In the bivariate logistic regression analysis; age, sex, residence, occupation, BMI, LoC, fruit consumption and past history of smoking had a statistically significant relationship with multimorbidity of chronic NCDs. However, only age, BMI and LoC were the variables that remain statistically significant in the adjusted model. In contrast to the existing evidence, wealth index (scored as a proxy for SES) did not show any likelihood of relationship with multimorbidity in this study (Table 5).

**Table 5:**
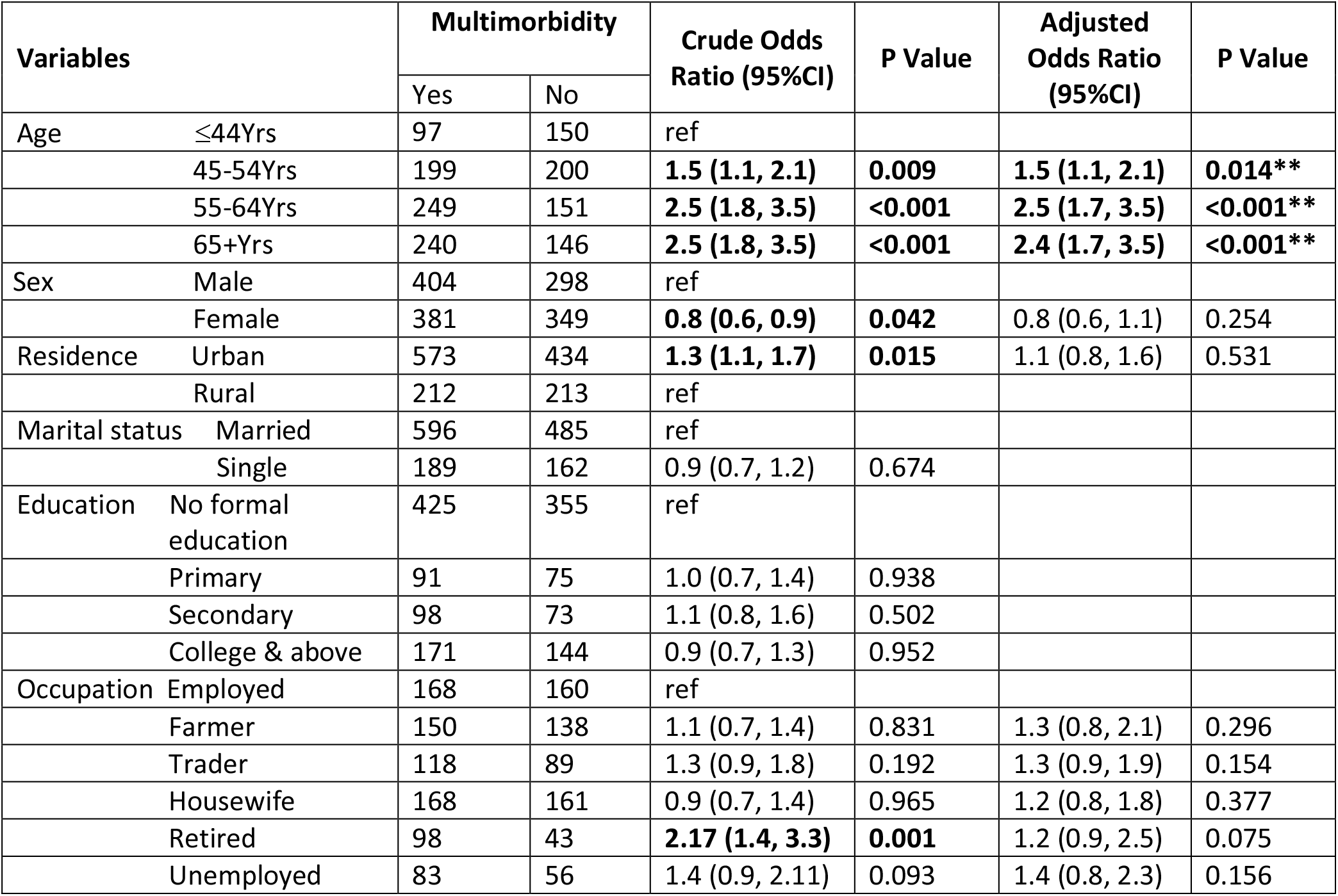
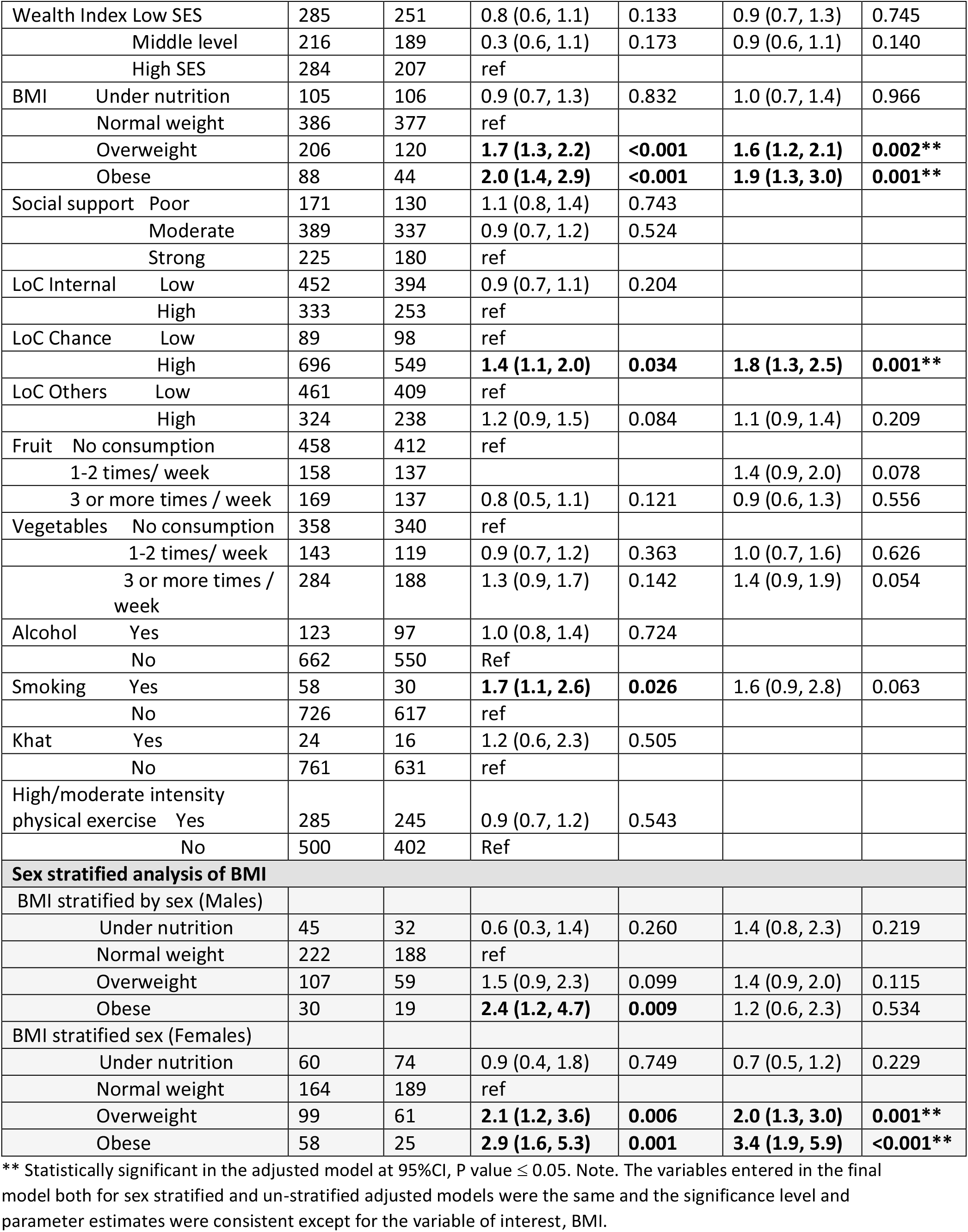
Bivariate and Multivariable Analysis of the Factors Associated with Multimorbidity, Bahir Dar, Ethiopia.

| Variables |  | Multimorbidity |  | Crude Odds Ratio (95%CI) | P Value | Adjusted Odds Ratio (95%CI) | P Value |
| --- | --- | --- | --- | --- | --- | --- | --- |
|  |  | Yes | No |  |  |  |  |
| Age | ≤44Yrs | 97 | 150 | ref |  |  |  |
|  | 45-54Yrs | 199 | 200 | **1.5 (1.1, 2.1)** | **0.009** | **1.5 (1.1, 2.1)** | **0.014\*\*** |
|  | 55-64Yrs | 249 | 151 | **2.5 (1.8, 3.5)** | **<0.001** | **2.5 (1.7, 3.5)** | **<0.001\*\*** |
|  | 65+Yrs | 240 | 146 | **2.5 (1.8, 3.5)** | **<0.001** | **2.4 (1.7, 3.5)** | **<0.001\*\*** |
| Sex | Male | 404 | 298 | ref |  |  |  |
|  | Female | 381 | 349 | **0.8 (0.6, 0.9)** | **0.042** | 0.8 (0.6, 1.1) | 0.254 |
| Residence | Urban | 573 | 434 | **1.3 (1.1, 1.7)** | **0.015** | 1.1 (0.8, 1.6) | 0.531 |
|  | Rural | 212 | 213 | ref |  |  |  |
| Marital status | Married | 596 | 485 | ref |  |  |  |
|  | Single | 189 | 162 | 0.9 (0.7, 1.2) | 0.674 |  |  |
| Education | No formal education | 425 | 355 | ref |  |  |  |
|  | Primary | 91 | 75 | 1.0 (0.7, 1.4) | 0.938 |  |  |
|  | Secondary | 98 | 73 | 1.1 (0.8, 1.6) | 0.502 |  |  |
|  | College & above | 171 | 144 | 0.9 (0.7, 1.3) | 0.952 |  |  |
| Occupation | Employed | 168 | 160 | ref |  |  |  |
|  | Farmer | 150 | 138 | 1.1 (0.7, 1.4) | 0.831 | 1.3 (0.8, 2.1) | 0.296 |
|  | Trader | 118 | 89 | 1.3 (0.9, 1.8) | 0.192 | 1.3 (0.9, 1.9) | 0.154 |
|  | Housewife | 168 | 161 | 0.9 (0.7, 1.4) | 0.965 | 1.2 (0.8, 1.8) | 0.377 |
|  | Retired | 98 | 43 | **2.17 (1.4, 3.3)** | **0.001** | 1.2 (0.9, 2.5) | 0.075 |
|  | Unemployed | 83 | 56 | 1.4 (0.9, 2.11) | 0.093 | 1.4 (0.8, 2.3) | 0.156 |

|  |  |  |  |  |  |  |
| --- | --- | --- | --- | --- | --- | --- |
| Wealth Index Low SES | 285 | 251 | 0.8 (0.6, 1.1) | 0.133 | 0.9 (0.7, 1.3) | 0.745 |
| Middle level | 216 | 189 | 0.3 (0.6, 1.1) | 0.173 | 0.9 (0.6, 1.1) | 0.140 |
| High SES | 284 | 207 | ref |  |  |  |
| BMI Under nutrition | 105 | 106 | 0.9 (0.7, 1.3) | 0.832 | 1.0 (0.7, 1.4) | 0.966 |
| Normal weight | 386 | 377 | ref |  |  |  |
| Overweight | 206 | 120 | **1.7 (1.3, 2.2)** | **<0.001** | **1.6 (1.2, 2.1)** | **0.002\*\*** |
| Obese | 88 | 44 | **2.0 (1.4, 2.9)** | **<0.001** | **1.9 (1.3, 3.0)** | **0.001\*\*** |
| Social support Poor | 171 | 130 | 1.1 (0.8, 1.4) | 0.743 |  |  |
| Moderate | 389 | 337 | 0.9 (0.7, 1.2) | 0.524 |  |  |
| Strong | 225 | 180 | ref |  |  |  |
| LoC Internal Low | 452 | 394 | 0.9 (0.7, 1.1) | 0.204 |  |  |
| High | 333 | 253 | ref |  |  |  |
| LoC Chance Low | 89 | 98 | ref |  |  |  |
| High | 696 | 549 | **1.4 (1.1, 2.0)** | **0.034** | **1.8 (1.3, 2.5)** | **0.001\*\*** |
| LoC Others Low | 461 | 409 | ref |  |  |  |
| High | 324 | 238 | 1.2 (0.9, 1.5) | 0.084 | 1.1 (0.9, 1.4) | 0.209 |
| Fruit No consumption | 458 | 412 | ref |  |  |  |
| 1-2 times/ week | 158 | 137 |  |  | 1.4 (0.9, 2.0) | 0.078 |
| 3 or more times / week | 169 | 137 | 0.8 (0.5, 1.1) | 0.121 | 0.9 (0.6, 1.3) | 0.556 |
| Vegetables No consumption | 358 | 340 | ref |  |  |  |
| 1-2 times/ week | 143 | 119 | 0.9 (0.7, 1.2) | 0.363 | 1.0 (0.7, 1.6) | 0.626 |
| 3 or more times / week | 284 | 188 | 1.3 (0.9, 1.7) | 0.142 | 1.4 (0.9, 1.9) | 0.054 |
| Alcohol Yes | 123 | 97 | 1.0 (0.8, 1.4) | 0.724 |  |  |
| No | 662 | 550 | Ref |  |  |  |
| Smoking Yes | 58 | 30 | **1.7 (1.1, 2.6)** | **0.026** | 1.6 (0.9, 2.8) | 0.063 |
| No | 726 | 617 | ref |  |  |  |
| Khat Yes | 24 | 16 | 1.2 (0.6, 2.3) | 0.505 |  |  |
| No | 761 | 631 | ref |  |  |  |
| High/moderate intensity physical exercise Yes | 285 | 245 | 0.9 (0.7, 1.2) | 0.543 |  |  |
| No | 500 | 402 | Ref |  |  |  |
| **Sex stratified analysis of BMI** |  |  |  |  |  |  |
| BMI stratified by sex (Males) |  |  |  |  |  |  |
| Under nutrition | 45 | 32 | 0.6 (0.3, 1.4) | 0.260 | 1.4 (0.8, 2.3) | 0.219 |
| Normal weight | 222 | 188 | ref |  |  |  |
| Overweight | 107 | 59 | 1.5 (0.9, 2.3) | 0.099 | 1.4 (0.9, 2.0) | 0.115 |
| Obese | 30 | 19 | **2.4 (1.2, 4.7)** | **0.009** | 1.2 (0.6, 2.3) | 0.534 |
| BMI stratified sex (Females) |  |  |  |  |  |  |
| Under nutrition | 60 | 74 | 0.9 (0.4, 1.8) | 0.749 | 0.7 (0.5, 1.2) | 0.229 |
| Normal weight | 164 | 189 | ref |  |  |  |
| Overweight | 99 | 61 | **2.1 (1.2, 3.6)** | **0.006** | **2.0 (1.3, 3.0)** | **0.001\*\*** |
| Obese | 58 | 25 | **2.9 (1.6, 5.3)** | **0.001** | **3.4 (1.9, 5.9)** | **<0.001\*\*** |

\*\* Statistically significant in the adjusted model at 95%CI, P value $\leq 0.05$ . Note. The variables entered in the final model both for sex stratified and un-stratified adjusted models were the same and the significance level and parameter estimates were consistent except for the variable of interest, BMI.

Compared to participants younger than 45 years, participants falling on the age group 45-54 years (AOR: 1.5, 95%CI= 1.1, 2.1), 55-64 years (AOR: 2.5, 95%CI=1.7, 3.5) and 65 years or more (AOR: 2.4, 95%CI=1.7, 3.5) had a statistically significant association with multimorbidity. Similarly, individuals classified as overweight (AOR: 1.6, 95%CI=1.2, 2.1) or obese (AOR: 1.9, 95%CI=1.3, 3.0) in the BMI category were 1.6 and 1.9 times more likely to have multimorbidity than those in the normal weight category, respectively. The sex stratified regression analyses revealed that, the effect of obesity on the likelihood of developing multimorbidity was implicated by female gender. We observed that women in the overweight and obese BMI category had 2 and 3.4 times higher odds of multimorbidity compared to the reference BMI category, respectively. However, obesity in males did not show any significant relationship with multimorbidity in the final model. In addition, individuals who strongly believe that life events and illnesses occur due to chance were 1.8 times more likely to have a-higher odds of multimorbidity than individuals who reported a lower rate of chance as a responsible factor for diseases to occur (AOR:1.8, 95%CI=1.3, 2.5) (Table 5).

### Fitting the LCA Model

One of the most important tasks in using LCA is accurately identifying the number of underlying latent classes and correctly placing individuals into their respective classes with a high degree of confidence(49).

We used R software to run the LCA model. Only chronic NCDs accounting 5% or more in the prevalence estimate were eligible for fitting in the LCA model (50). The diseases satisfying the inclusion criteria were hypertension, diabetes, heart diseases, stroke, depression and asthma (scored as yes or no value). While fitting the LCA model, we employed a sequential process (48), started with a one-class model and then added one class at a time and continued to run models with one additional class at a time until the best model is identified. We compared and identified the best model based on statistical and theoretical criteria.

Fit statistics, including the Akaike information criterion (AIC), Bayesian information criterion (BIC), the likelihood ratio, chi-square goodness of fit and entropy were compared and the class model with the lowest BIC has been chosen as the best model (48). The BIC is destined to be the most reliable fit statistic to evaluate model fit and the lower the BIC value of the statistic the more efficient the model is to fit the data (49). In our analysis, the four class model had the lowest BIC value and the highest entropy and the addition of classes beyond four provides essentially no improvement in fit (Table 6).

**Table 6:** Model fit statistics of each class fitted in the LCA model, Bahir Dar, Ethiopia.

| Latent class model | AIC | BIC | Entropy | Gsq | Chisq |
| --- | --- | --- | --- | --- | --- |
| 1 class model | 7835.99 | 7867.58 | ----- | 688.63 | 613.86 |
| 2 class model | 7499.47 | 7567.94 | 0.80 | 338.13 | 311.90 |
| 3 class model | 7342.26 | 7447.34 | 0.72 | 166.67 | 174.05 |
| 4 class model | 7261.77 | **7402.97** | **0.86** | 71.43 | 66.69 |
| 5 class model | 7255.77 | 7434.84 | 0.67 | 52.42 | 52.43 |
| 6 class model | 7244.18 | 7460.12 | 0.70 | 26.83 | 24.62 |

Note. Gsq=the likelihood-ratio statistic; Chisq=Pearson Chi-square goodness of fit statistic; Entropy=the extent to which the groups identified are distinct from one another (its value ranges between 0 and 1 and a value $\geq 0.8$ shows good separation of the group).

Based on the average latent class posterior probability (49), the four class model classified 50.2% of the participants in class 1, 32.6% of the participants in class 2, 11.5% of the participants in class 4 and the remaining 5.7% in class 3 (Figure 3).

**Figure 3:**
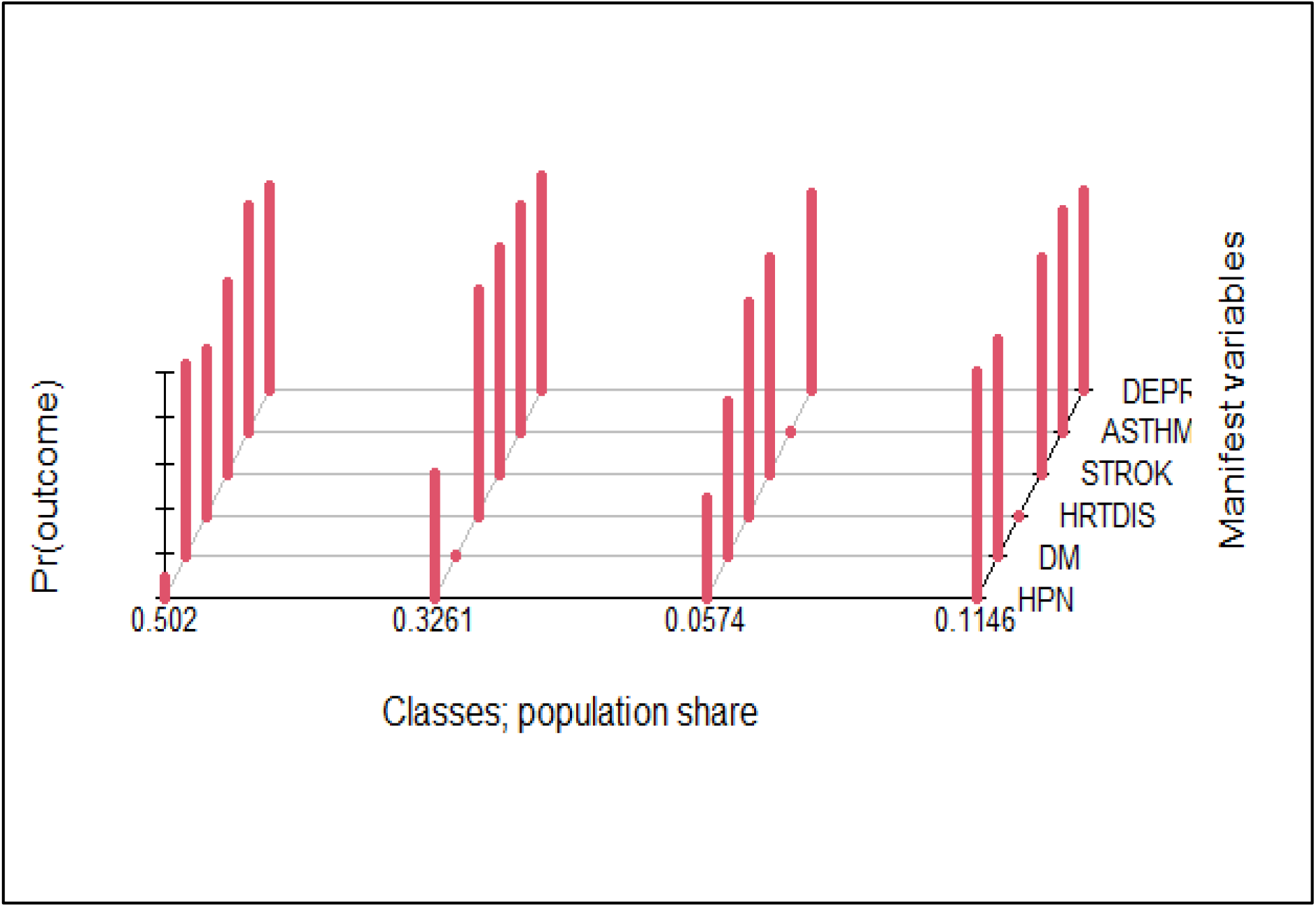
Population share and distribution of conditions in the latent classes, Bahir Dar, Ethiopia.

Upon further analysis, although not deterministic, the model predicted the probability of belongingness of a person having a “yes” response to each indication variables. For example, a person having hypertension was a 100% probability of being assigned in class 1, 44% of being in class 4 and 11% in class 3 and 0% in class 2. Those having diabetes had a 100% probability of being assigned in class 4, 16% of being in class 1, 6% of being in class 2 and 4% of being in class 3. Looking into those reported to have heart diseases, the model predicted 100% of their being in class 2, 26% in class 1 and 0% in class 3 and 4. The probability of being in class 3 for those reported to have asthma, stroke and depression and was 42, 36% and 19%, respectively (Table 7).

**Table 7:** The predicted probability that a person responding “yes” to the indicator variables is being assigned in the classes in the LCA model, Bahir Dar, Ethiopia.

| Variables | Predicted Probability |  |  |  |
| --- | --- | --- | --- | --- |
|  | Latent Classes |  |  |  |
|  | Class 1 | Class 2 | Class 3 | Class 4 |
| Hypertension | 1.00 | 0.00 | 0.11 | 0.44 |
| Diabetes | 0.16 | 0.06 | 0.04 | 1.00 |
| Heart diseases | 0.26 | 1.00 | 0.00 | 0.00 |
| Stroke | 0.12 | 0.06 | 0.36 | 0.01 |
| Depression | 0.09 | 0.11 | 0.19 | 0.05 |
| Asthma | 0.05 | 0.03 | 0.42 | 0.04 |
| Latent class marginal posterior probabilities | 0.52 | 0.21 | 0.05 | 0.22 |
| **Class size** | **622** | **184** | **68** | **558** |

As the observed sample is a mixture of individuals from different latent classes, individuals belonging to the same class are similar to one another such that their observed scores on a set of indicators are assumed to come from the same probability distribution(48, 49). Hence, according to the distribution of individuals responding yes to one or more of the indicator variables, class 1 contains majority of the individuals with hypertension, about half of the individuals with heart diseases and majority of individuals with stroke, and named as a class with cardiovascular pattern. Class 2 contains majority of individuals with heart diseases and one third of individuals with depression and hence names as a class with cardio-mental pattern. Whereas, class 4 contains the large majority of individuals with diabetes and nearly one third of individuals with hypertension and named as metabolic patterns and class 3 contains higher proportion of individual with asthma and small proportion of individuals with stroke and named as a class with respiratory pattern. Although the class separation seems adequate and the 4 class LCA model is parsimonious, individuals might not be placed adequately in a discrete and non-overlapping way.

As shown on table 8, none of the classes had a class size of less than 50. While class 1 includes the majority (n=622, 43.4%), class 3 had the lowest number (n=68, 4.7%) of individuals.

**Table 8:**
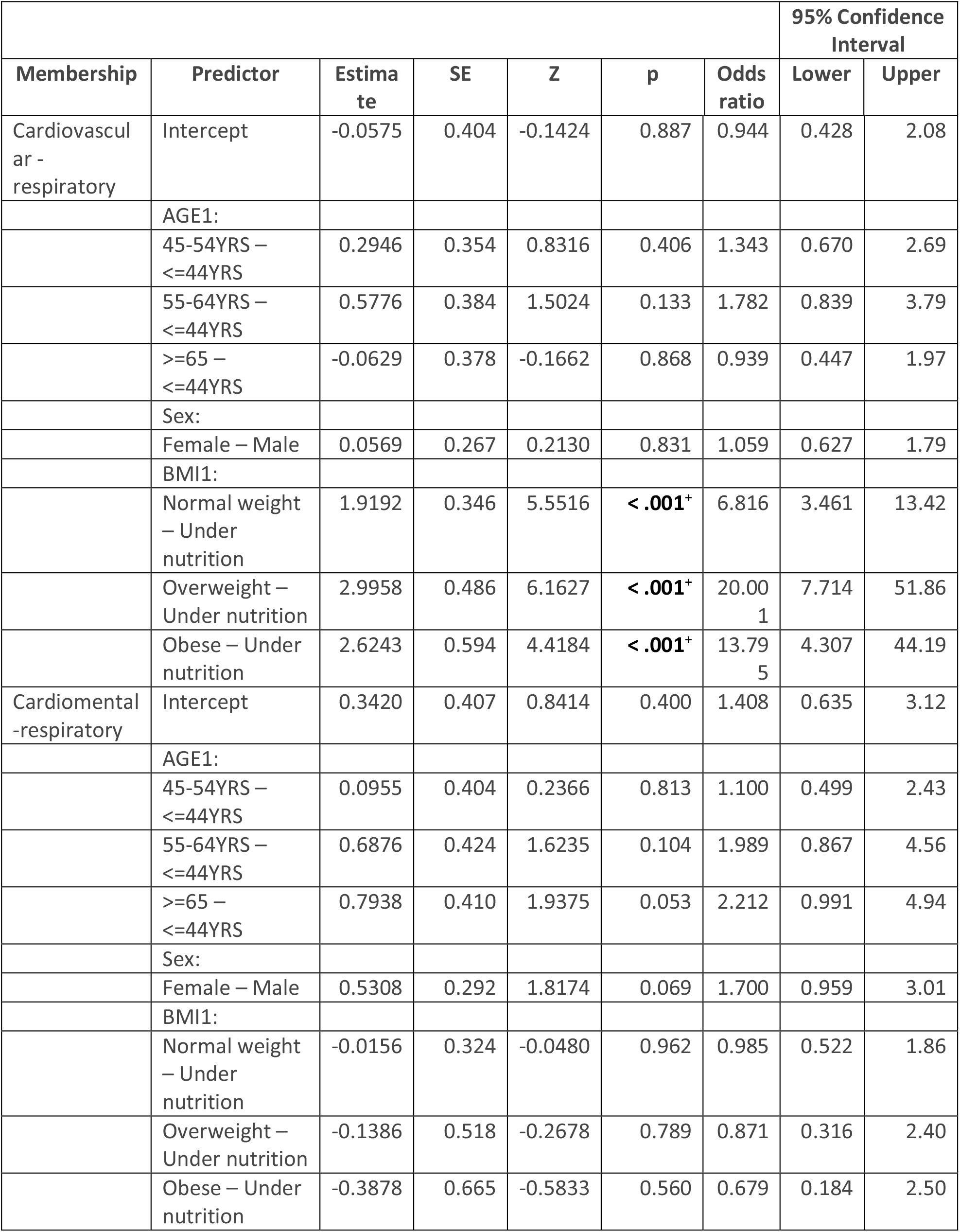
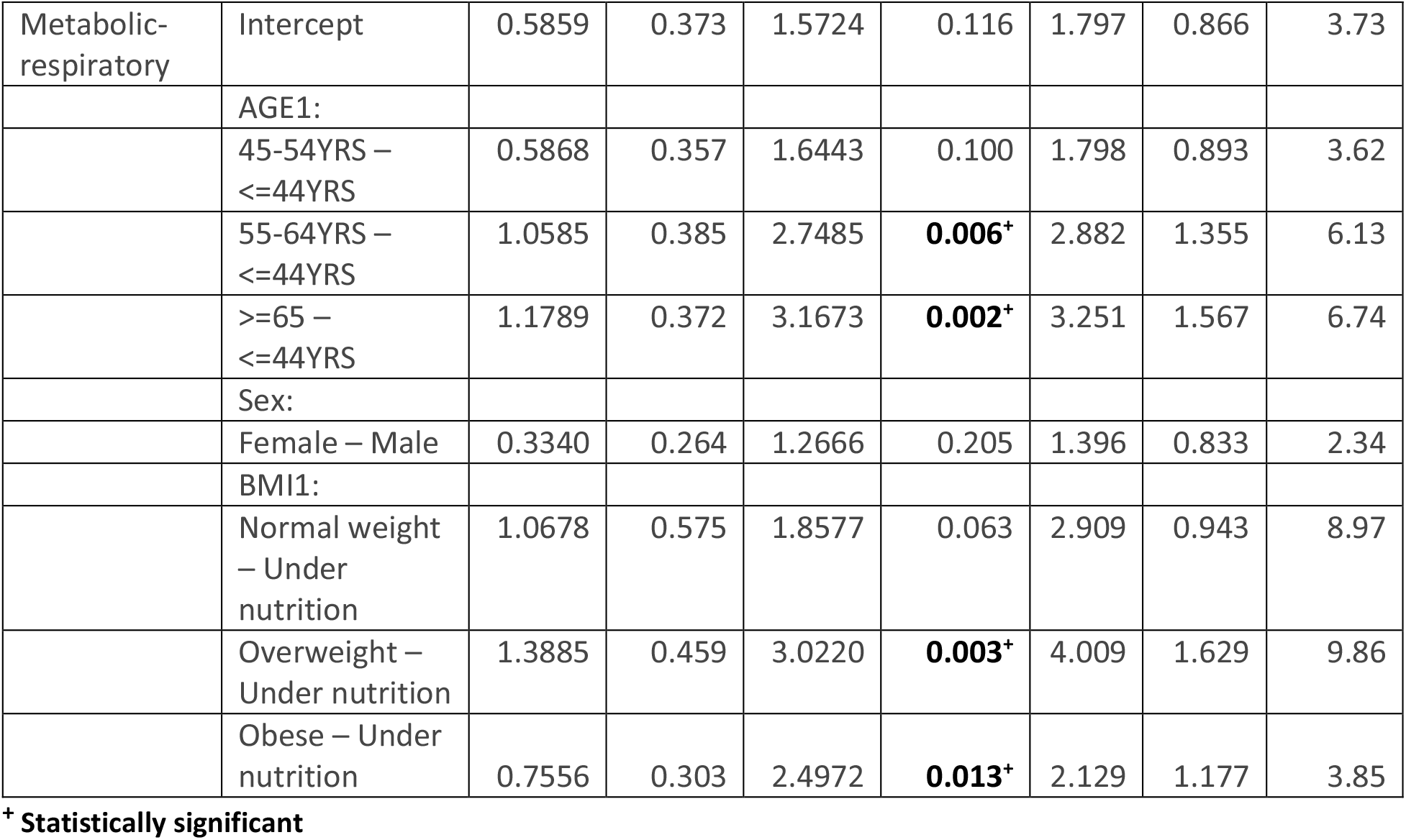
Multinomial logistic regression analysis of the factors determining latent class membership, Bahir Dar, Ethiopia.

### Predictors of Class Membership in the LCA Model

The names we provided to categorize latent classes (patterns of distribution) were cardiovascular, cardio mental, respiratory and metabolic. We run multinomial logistic regression analysis using these categories as dependent variables and by making the respiratory group as a reference category. We determined the effect of sex, age group and BMI level of individuals in predicting class membership. The model we fitted (in SPSS) satisfied the significance in the likelihood ratio of model fitness and classification accuracy criteria (predicted estimate of the group membership was higher (49.7%) than the actual group membership value (44.9%)). Furthermore, as multinomial regression provides unbiased estimates when the data meet the independence of irrelevant alternatives (IIA) assumption (Hausman test of significance) (in STATA) (51), we run both the fixed effect and random effect test and checked the significance level in the Hausman test output. The output showed a Chi-square value of 12.59 and p-value 0.0018, signifying to reject the null hypothesis and use the fixed effect model rather than the random effect model(51).

As shown on table 9 below, the implication of BMI and age in dictating class membership was profound. The odds of being classified in the cardiovascular group rather than the respiratory group for individuals classified as normal weight, overweight and obese was 6.8, 20.0 and 13.8, respectively times higher than those classified in the underweight group. On the other hand, compared to individuals grouped in the respiratory class, the odds of being classified in the metabolic group were 2.8 and 3.2 times higher for individuals in the age group 55-64 years and 65+years, respectively than those individuals in the reference category (≤44 years). Similarly, as opposed to those classified in the respiratory class, individuals classified as overweight and obese were 4.0 and 2.1 times more likely to be classified in the metabolic group than those classified in the underweight group. Except for the metabolic group, age did not predict class membership of individuals in our analysis. Further, we did not observe any statistically significant effect of gender in predicting class membership in the multinomial logistic regression model (Table 8).

## Discussion

This study investigated the magnitude and patterns of multimorbidity, factors associated with multimorbidity and the underlying latent classes of individuals behind a set of indicators variables (chronic NCDs). Understanding the extent and patterns of NCDs combination could inform design of effective intervention owing to the prevention and management of multimorbidity. We employed a blend of methods to accurately measure the burden of each individual NCD and their pairwise and triple combination among a broad sample of 1432 individuals (aged 40+) attending chronic care in hospitals and specialized health facilities in Bahir Dar city, northwest Ethiopia.

The magnitude of multimorbidity estimated in this study was high (55%) and a significant proportion (15.2%) of the participants had three or more chronic conditions. A striking toll of multimorbidity among the people on chronic care follow-up in this study would mean multimorbidity has been and will continue to posing a huge burden on individuals, health systems and the society at large. This finding is in agreement with what is abundant in the global literature that proclaims the emergence of multimorbidity as “a norm than the exception” both in high-income and low-and middle-income countries (18).

Consistent with previous studies (52), hypertension remain the leading NCD affecting 65% of the people visiting health facilities in the study area. Perhaps, this is a reflection that hypertension may be burdensome in the society. Diabetes was the second highly reported chronic condition among respondent and heart diseases took the third rank among the reported conditions in the patient population. These diseases played a significant role in shaping the profile of disease combinations and patterns of multimorbidity in this study. The significance of hypertension, diabetes and heart diseases in dictating patterns of multimorbidity has also been reported elsewhere (9, 53, 54).

In agreement with several studies in LMICs (52, 55), the most prevalent pairwise combination was between hypertension and diabetes. Hypertension was also frequently comorbid with heart diseases. Such peculiarities in combination of NCDs in the study area may substantiate the probability that these diseases may have shared risk factors or have a common pathway of occurrence or there is a causal relationship between them(48). The pattern of combinations of conditions identified in the observational clustering was almost consistent with the taxonomy assigned based on the class membership in the LCA model.

Among the four distinctive groups of individuals in the sample, the largest disease burden was seen in cardiovascular (hypertension + heart diseases) cluster, followed by metabolic (diabetes + hypertension) cluster, which together comprised 82.4% of the pattern. Whereas, the cardio-mental and respiratory group comprise 11.5% and 5.7%, respectively.

Two of the risk factors identified for the development of multimorbidity; age group and BMI have also significantly contributed for determining group membership. Although it may be difficult to directly compare our findings with those of the previous studies owing to the differences in the underlying demographic characteristics, disease profiles, sources of data and study setting, the patterns of NCDs distribution and pattern of multimorbidity observed in our study concur with what is reported in some previous studies(13).

Individuals living with different types and combinations of NCDs may have different needs and priorities(56). Perhaps living with a similar pattern of combination would have different effect on different individuals. Therefore, health systems need to account this variability as the notion of one-size-fits-all will not be helpful anymore (13).

### Factors associated with multimorbidity

Advanced age is always been associated with occurrences of multimorbidity globally (1, 57, 58). Evidence shows multimorbidity starts at around 40 years of age and continue to increase in prevalence until 70 years and remain constant after then owing to the reduced survival rate of those people aged 70+ (18). In this study, we observed an increasing and statistically significant likelihood of multimorbidity among individuals aged 45+. The higher odds of multimorbidity among the people aged 55-64 and 65+ implies that longevity plays a profound role in rising the incidences of multiple chronic conditions and its impact on quality of life and functioning (1, 18). However, studies in high income countries reported that people living in socioeconomically deprived areas tend to develop multimorbidity 10-15 years earlier than their wealthier counterparts (1), implying that income could modify the onset of multimorbidity in low income setting.

The relationship between unhealthy diet, obesity and overweight and multimorbidity is well established globally (57, 59, 60). In this study, individuals classified as obese and overweight were more likely to have higher odds of multimorbidity than individuals with normal weight. Globally, obesity is a known risk factor for the occurrences of NCDs (61) and much of the increase in multimorbidity among the people living with NCDs was due to obesity (62, 63). Studies found obese individuals particularly women, Gen Xers and younger boomers have a higher risk of multimorbidity than those of normal weight (64). In consistent with existing evidence, the probability of living with multimorbidity was high among women identified as overweight and obese compared to males in the same category upon running a sex stratified analysis in our study.

individuals with external LoC tend to ascribe their circumstances to the effects of outside factors, not under their personal control (57, 65). In this study, individuals having a higher level of believing in the attribute of chance in causing diseases shown to have increased probability of multimorbidity. This finding is in line with the knowledge established in this regard globally. The evidence base in establishing a causal relationship between LoC and multimorbidity is not conclusive, however (66).

The lack of association with female sex is not new; previous findings are inconsistent on the association between gender and multimorbidity (57). However, the lack of association of SES with multimorbidity in our study might be due the nature of data we collected to estimate wealth index in the context. Information on the types of materials respondents’ houses were constructed, ownership of material assets and livestock, household’s water sources and toilet facilities, cash crop production and ownership of agricultural and non-agricultural lands were the main variables measured and considered for estimating household income using the principal component analysis (PCA) separately for urban and rural residents.

We did not find any statistically significant association of dietary habits and behavioral practices such as physical exercise, alcohol intake and smoking with multimorbidity. Although evidence is not conclusive globally (13, 67), the lack of association between these variable and multimorbidity in our study may not only be due to lack of causal relationship per se, but might also be due to the potential of recall and social desirability biases introduced during data collection.

### Implication for Research, Policy and Practice

Multimorbidity is a priority global health research agenda (68). However, the methodologies employed to study the epidemiology of multimorbidity are not consistent globally (7, 54, 69). A number of studies argue that estimates of multimorbidity may vary depending on the age of the population studied, the type and number of chronic conditions considered in determining multimorbidity, study settings (population based vs facility based) where multimorbidity researches are conducted and the sources of data used to identify presence of chronic conditions (6, 70).

There is a huge variability in the age group of individuals enrolled in multimorbidity studies globally (71). The mean age of individuals participated across most of facility based multimorbidity studies in LMICs ranged from 40 -45 years (9, 52). To facilitate comparison and because of the increased focus of studying multimorbidity among middle-aged and elderly people (3, 72), we only enrolled participants aged 40 years or more. Future studies may, however, investigate the distribution of individual NCDs and their patterns of combination in a broad age category.

Compared to population based studies, facility based studies foster a wealth of data sources to validate and triangulate information about presence of individual NCDs and other peculiarities (6). We used two complimentary methods (interview of a doctor diagnosed condition and medical records review) to better estimate the magnitude of individual conditions. The type of chronic NCDs included in this study was selected based on a previous study (9) and analysis of preliminary data collected prior to the actual study. The magnitude of individual conditions and the multimorbidity estimated in this study were higher due to the involvement of patients on follow-up for any of the 13 selected chronic NCDs. However, the estimate in our study may not necessarily reflect the underlying epidemiology in the general population. Therefore, researchers need to be aware of these limitations when considering facility based multimorbidity studies.

Evidence (73) has shown that population based studies are better in producing prevalence estimates. However, most population based studies were limited to self-report (simple count) data as employing diagnostic methods and technologies to identify a broad range of conditions is not feasible, particularly in low income settings(71). Studies relying on self-report data alone tend to over or under estimate prevalence rates. The importance of using weighted multimorbidity measures such as the Charlson index (74) and multimorbidity index(75) has been highlighted in studying the burden and outcomes of multimorbidity.

The number of chronic conditions to be considered for measuring multimorbidity has been one of the areas of debate globally (6, 66). The list of chronic condition among studies in LMICs ranged from 3-40 (9), leading to inconsistent results. Prevalence rates considering 4 to 7 diagnoses or fewer than 10 chronic conditions may lead to an underestimation of the prevalence of multimorbidity (6, 76). To better estimate multimorbidity, it is recommended to consider a minimum threshold of 12 chronic conditions that have public health importance in a given context (6). We used 13 most prevalent chronic NCDs (including those grouped together) in the study area. However, scholars in the field (48, 72) emphasized the importance of also evaluating the severity of the disease conditions beyond a simple count of combination of medical diagnoses. Others also suggest the inclusion of HIV (when prevalence is high), obesity and high cholesterol in the list to define multimorbidity (73). Any list of conditions in the pursuit of operationalizing multimorbidity should at least consider disease burden (prevalence) and their impact on population wellbeing and survival in a given context (68, 77).

It is argued that patients with multimorbidity are more than the sum of their individual conditions (13, 78) and that, the patterns of disease combination and severity level influence the health care to be delivered and the subsequent outcomes (78, 79). The 4 class LCA model was parsimonious to identify common multimorbidity patterns and the covariates determining class membership. Individuals belonging to the same patterns of classification are similar one another(13). For example, a patient suffering from two conditions, such as stroke and heart failure does not have the same burden of illness as does a patient with hypertension and diabetes (80). Hence, it is imperative to define the best possible model of health care for people with different patterns of multimorbidity(81).

Although evidence is mixed and inconclusive, the notion of integrated and person-centered care and coordination in the pathway and continuity of care over time have been underscored (82). In addition, multimorbidity should drive a shift in the way health policies are developed and guide the health care system in tackling this challenge. In this sense, it is clear that priority should be directed to develop clinical practice guidelines and strengthen the primary care services. The development of clinical practice guidelines should fuel a reform in the academic curriculum and continuing training programs to accommodate the new scenario in health professions’ education (9).

### Strengthen and Limitation of the study

Our study has the advantage of including a broader range of health care facilities providing comprehensive health services for the people living with NCDs. Guided by a published study protocol, this study employed two complimentary methods to accurately identify the diagnosed chronic NCDs. The LCA model we fitted helped to plausibly determine the pattern of multimorbidity and identify covariates influencing class membership in a relatively efficient, reliable and valid way.

However, the findings of this facility based study may not exactly represent the underlying epidemiology and patterns of distribution of NCDs in the population. However, understanding the nature of people visiting health facilities, the types and magnitude of NCDs they are seeking care for and patterns of combination of NCDs among the cohort of individuals in our study would help guide resource planning, prevention and management of multimorbidity in the country.

### Conclusion and Recommendation

The magnitude of multimorbidity in this study was high. The most frequently diagnosed chronic conditions shaped the profiles of clustering and patterns of multimorbidity. The proportion of individuals living with three more chronic NCDs was also significant. Advanced age, overweight and obesity in females and external locus of control were the factors associated with multimorbidity. The high multimorbidity estimate observed in this study might be attributed to the fact that the study was conducted among health facilities where most of people living with chronic NCDs were attending care.

The literature on multimorbidity is dominated by studies in high income countries. If health systems in LMICs are to meet the challenges of multimorbidity, we need to first investigate its magnitude, associated factors and the pattern of disease clustering. We employed more sensitive methods to capture the full breadth of multimorbidity across the ages and the patterns identified should help paving the way in designing and rendering comprehensive and patient center care.

Health service organization and provision in the study area need to be configured owing to the highest proportion of the people with distinctive nature of comorbidity such as the coexistence between hypertension and diabetes, and hypertension and heart diseases.

Future studies may need to involve people with broader age groups such as including adults younger than 40 years. Although recall bias, diagnostic uncertainty and resource constraint may hinder implementation of population base studies in our setting, understanding the epidemiology of multimorbidity in the population is imperative to confirm our tentative conclusion that the trend we observed in our study may be the reflection of the underlying phenomenon in the society. Further studies (such as the one we are conducting) are also needed to explore the effect of multimorbidity on quality of life, functioning and survival and assess how health services are oriented and organized to meet the care needs of the people living multiple chronic conditions.

### Ethics approval and consent to participate in the study

As this is a part of an ongoing PhD study, permission to conducting the study has been obtained from the Institutional Review Board (IRB) of the college of medicine and health sciences, Bahir Dar University with a protocol number 003/2021. Study participants were enrolled after consenting to participate in the study. Permission was obtained from the health facilities involved in the study. Moreover, confidentiality of the data we obtained from study participants and medical records have been strictly maintained.

## Data Availability

All data produced in the present work are contained in the manuscript

## Acknowledgements

We thank Bahir Dar University for partially funding this study. We also thank Jhpiego-Ethiopia for the facilities we used to conduct this study. We would like to also thank data collectors, supervisors, facilities leaders and study participants for their support in making this study a reality.

## Author Contributions

FAE, FAG, MS and SA conceived and designed this study. FAE, FAG and MS participated in the data analysis and interpretation of the findings. FAE wrote the manuscript and SP contributed in revising the manuscript. All authors critically reviewed and approved the final manuscript for submission.

## Funding statement

### Funding

This work was partially supported by Bahir Dar University [grant number: RCS/003/21].

### Disclaimer

The funders had no roles in study design, fieldwork, data analysis, interpretation, and decision to publish the manuscript.

### Competing interests

None declared.

### Patient consent for publication

Not required.

### Data Availability Statement

in compliance with PLOS’ data policy, all relevant data are included in the article and will also be published in relevant repositories accordingly.

## Notes

### Competing Interest Statement

The authors have declared no competing interest.

### Funding Statement

This study was partially funded by Bahir Dar university

### Author Declarations

This study was approved by Institutional Review Board (IRB) of the college of medicine and health sciences, Bahir Dar University with a protocol number 003/2021.

